# Patterns of early neocortical amyloid beta accumulation: a positron emission tomography population-based study

**DOI:** 10.1101/2023.05.14.23289948

**Authors:** Emily E. Lecy, Hoon-Ki Min, Christopher J. Apgar, Daniela D. Maltais, Emily S. Lundt, Sabrina M. Albertson, Matthew L. Senjem, Christopher G. Schwarz, Hugo Botha, Jonathan Graff-Radford, David T. Jones, Prashanthi Vemuri, Kejal Kantarci, David S. Knopman, Ronald C. Petersen, Clifford R. Jack, Jeyeon Lee, Val J. Lowe

## Abstract

**Introduction:** The widespread deposition of amyloid beta (Aβ) plaques in late-stage Alzheimer’s disease (AD) is well defined and confirmed by *in vivo* positron emission tomography (PET). However, there are discrepancies between which regions contribute to the earliest topographical Aβ deposition within the neocortex.

**Methods:** This study investigated Aβ signals in the peri-threshold SUVr range using Pittsburgh compound B (PiB) PET in a population-based study cross-sectionally and longitudinally. PiB-PET scans from 1,088 participants were assessed to determine the early patterns of PiB loading in the neocortex.

**Results:** Early-stage Aβ loading is seen first in the temporal, cingulate, and occipital regions. Regional early deposition patterns are similar in both Apolipoprotein ε4 (APOE) carriers and non-carriers. Hierarchical clustering analysis shows groups with different patterns of early amyloid deposition.

**Discussion:** These finding of initial Aβ deposition patterns may be of significance for diagnostics and understanding the development of different AD phenotypes.

## 1. INTRODUCTION

The neuropathology of Alzheimer’s disease (AD) is characterized by the deposition of Amyloid beta plaques (Aβ).[1, 2] Positron emission tomography (PET) using Aβ tracers has added to our understanding of Aβ deposition and AD progression. The first Aβ radiotracer, Pittsburgh Compound B (PiB), has been used in AD studies for more than a decade[3] and aligns with histological findings of Aβ localization.[4] Other Aβ PET biomarkers are currently available[5] and have been shown to have diagnostic accuracy similar to that of PiB, further establishing its efficacy.[6, 7] Currently, the widespread aggregation of Aβ plaques in late stage AD is well established;[8, 9] however, there are discrepancies across studies in how and where Aβ deposition begins.[10, 11]

Neuropathological studies describe the progression of Aβ deposition in ordered stages termed “Thal phases” in which deposition occurs in five phases.[12] The first Thal phase of isocortical Aβ deposition is defined as occurring exclusively in the neocortex, with exception of the paracentral lobule.[12] This cortical Aβ deposition is described as being diffusely distributed and without a specific neocortical regional pattern. While these postmortem histological studies provide conclusive results on the location of Aβ proteins at death,[13] it remains difficult to observe Aβ early progression because the majority of samples are from those whose Aβ onset was likely years prior and only 51 participants were evaluated.[12]

PET imaging studies provide a more detailed picture of neocortical deposition and longitudinal development *in vivo*. Past PET studies have analyzed different Aβ radiotracers and suggest areas where Aβ deposition begins; however, these studies show some inconsistencies in which regions early Aβ aggregation begins. Some describe ear-ly Aβ aggregation occurring in frontal areas such as frontotemporal association cortices,[14] frontomedial areas,[15] large-scale brain networks such as the default mode network (DMN),[16] parietal regions such as the precuneus,[15, 17] cingulate,[17] and medial orbitofrontal areas.[15, 17] There are discrepancies in the descriptions the temporal lobe in initial accumulation as some publications claim this to be a later aggregation point[15] while others deem it an early accumulation site.[14] Unfortunately, most of these studies have limitations by using pre-selected cohorts that limit the ability to generalize their results to the general population. Some do not assess the effect of risks factors on Aβ aggregation patterns, such as Apolipoprotein ε4 (APOE) status or familial history.[18] These inconsistencies in study design and conclusions of early aggregation of Aβ demonstrate a need to revisit the earliest patterns of Aβ in a population-based study.

In this work, PiB-PET is used in an epidemiological community-based population study to assess the prevalence of focal early Aβ signal changes across varying brain regions in the neocortex both cross-sectionally and longitudinally. To see subtle differences in Aβ deposition we: (1) selected participants who had an amyloid signal near the global PiB cut-off point,[19] called the Early PiB group, (2) determined elevated Aβ status for each ROI independently compared to younger cognitively unimpaired (CU) individuals [20] and (3) analyzed the elevated PiB data by ROI-wise analysis. Patterns of early regional Aβ deposition were assessed and cluster analysis was used to determine subgroups with different Aβ deposition patterns within the population.

## 2. MATERIALS AND METHODS

### 2.1 Participants

All chosen participants were enrolled in the Mayo Clinic Study of Aging (MCSA), a population-based randomized aging study from Olmsted County Minnesota of a wide age range.[21] Participants provided written consent with approval of Mayo Clinic and Olmsted Medical Center Institutional Review Boards. MCSA participants were invited to participate in imaging studies if they did not have contraindications. At enrollment and for all subsequent visits participants were clinically diagnosed as CU, having mild cognitive impairment (MCI), or having dementia via a consensus conference process (Supplementary Table 1).[22]

### 2.2 Neuroimaging and image analysis

Participants undergoing PiB-PET scans received a PiB dose (range 293.8-746.3 MBq), followed by a 33.5-64.5-minute post injection period. After, PET acquisition was taken for twenty minutes as previously described.[23] Cortical regions of interest (ROI) were defined by an in-house version of the automated anatomic labelling atlas[24] as previously described.[25] Standardized uptake value ratio (SUVr) image was calculated by dividing the median of uptake in the cerebellar crus grey matter. Regional SUVr uptake was defined as the median uptake across all grey matter voxels in a ROI. Two-component partial volume correction was used.[26] Global cortical PiB-PET SUVr was computed from a meta-region of interest.[19] Data may be available from the authors upon reasonable request and with permission.

### 2.3 Early PiB group and subgroups

To create a sample population that would be the most likely to have Aβ deposition, selected subjects that had amyloid signal near the global PiB cut-off point (SUVr of 1.42).[19] Thus, participants of this study, deemed the “Early PiB” group, were chosen if they were 50 years of age or older (50+), and had a global SUVr between 1.29 to 1.64 (Figure 1). The lower cut-off point in this range (1.29) was selected as the lower tertile global SUVr boundary of those 50+ in the MSCA who were CU. The upper limit of this range (1.64) was selected as the lower tertile boundary of the global SUVr for those 50+ in the MSCA with elevated amyloid levels. The Early PiB group (n=1,088) was comprised of 89.6% CU, 9.9% MCI, and 0.6% dementia (Supplementary Table 1).

**Figure 1.**
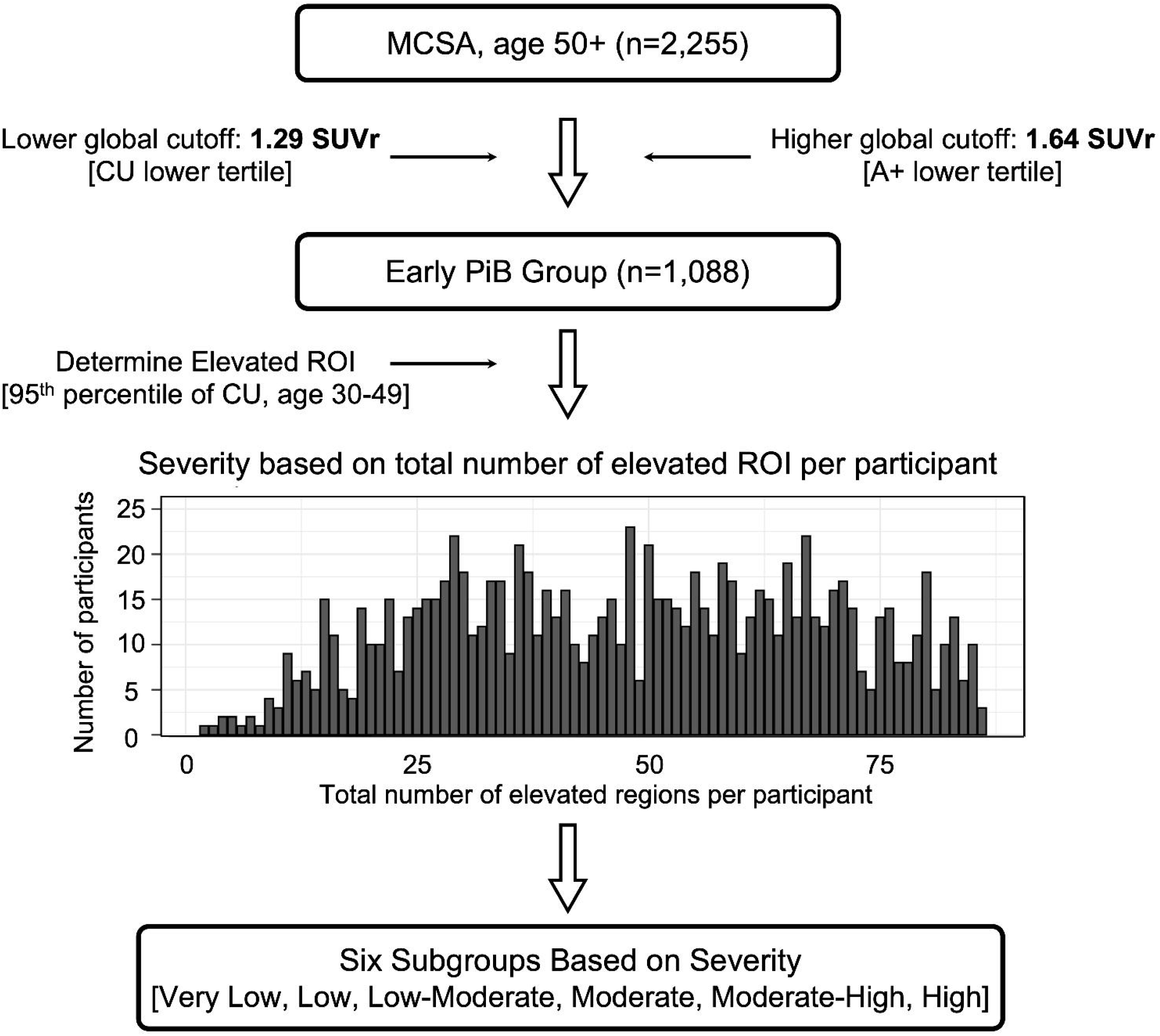
Participant selection criteria and regional PiB signal elevation by sub-grouping. Participants were selected from the Mayo Clinic Study of Aging (MCSA). For the Early PiB group, a specific SUVr cut point range created our population. The lower cut,1.29, was determined by the lower tertile of those who are cognitively unimpaired (CU) and of age 50 and above (50+) in the MCSA. The upper cut, 1.64, was determined by the lower tertile of those with global elevated amyloid (A+) and 50+ in the MCSA. A total of 1,088 participants fell within this range and are defined as the Early PiB group for this study. This Early PiB group was then distributed based on the number of total brain regions they had which presented with increased amyloid. People towards the right of the bar graph have multiple elevated PiB brain regions and those towards the left have fewer elevated brain regions, showing overall severity of PiB deposition. This distributed Early PiB group was further made into six equitably sized subgroups (very low, low, low-moderate, moderate, moderate-high, and high) based on the total number of brain regions presenting elevated PiB. Elevated PiB was determined by a region of interest (ROI) specific SUVr cut point derived from younger cognitively unimpaired individuals from the MCSA (30-49 years, n=164; Table 1).

The Early PiB group was then further distributed into subgroups of participants based on how many individual ROIs with elevated PiB levels were seen in each participant (i.e., the more elevated ROIs, the higher the participant group assignment). The regional elevated PiB level was determined by using region-specific cut-offs as being above the 95th percentile of younger CU MCSA individuals (30-49 years, n=146).[20] Groups were then defined with equitably participant-sized subgroupings. In all, six elevated ROI-based subgroups were created: very-low (n=170), low (n=180), low-moderate (n=185), moderate (n=186), moderate-high (n=190), and high (n=177) (where n=1,088, the respective participant number included). (Table 1, Figure 1, histogram).

### 2.4 SUVr based clustering analysis

Agglomerative hierarchical clustering analysis[27] with the Ward linkage method was performed using regional SUVr values (averaged over left and right hemispheres). We used Euclidean distance as a similarity measure. This iterative bottom-up algorithm combines pairs of clusters at each step while minimizing the sum of squared errors from the cluster mean. The number of clusters was fixed to 3 (K=3) a priori. The algorithm does not guarantee finding the optimal solution, and thus we also performed a k-means clustering analysis to compare the results.[28] Squared Euclidean distance was used as the similarity measure. The algorithm returns the K centroids maximizing intra-cluster similarity and maximizing inter-cluster dissimilarity. To compare rates of amyloid deposition by cluster we computed annualized percentage changes in SUVr for each cortical regions. Analyses were performed using R Statistical Software (version 3.6.2). The 3D volume rendering illustrations were created using the Surf Ice software (https://www.nitrc.org/projects/surfice/).

## 3. RESULTS

### 3.1 Cross-sectional stating of regional amyloid deposition

Elevated PiB-PET determined by region-specific cutoffs was observed in over 80% of Early PiB participants within the fusiform, angular gyrus, inferior and middle temporal, middle occipital, and calcarine region (Figure 2A). The amygdala and superior temporal pole had minimal elevation in PiB-PET SUVr, with elevation in under 25% of the population. The overall pattern of frequencies of amyloid-positivity was not visually different when applying the hemisphere specific-cutoff (left or right) or global hemispheric cutoff (voxel weighted median of left and right).

**Figure 2.**
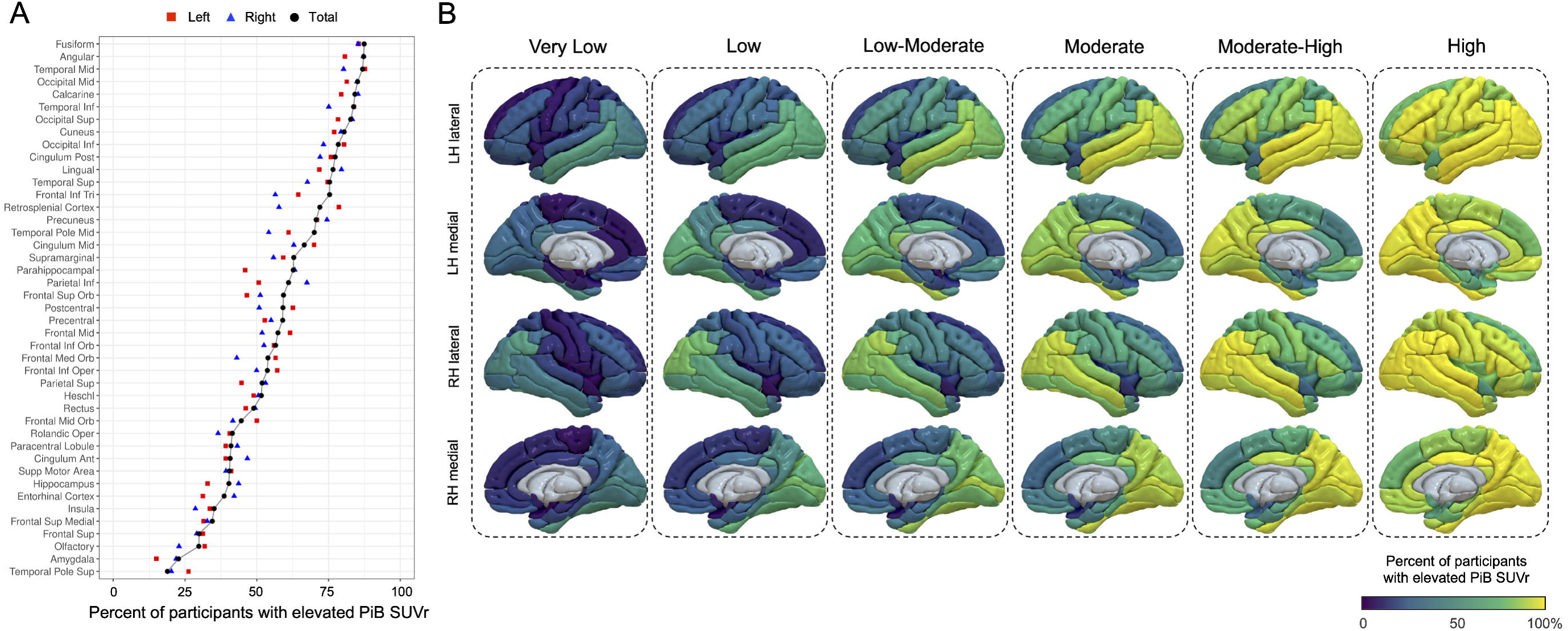
Percent of participants with elevated PiB PET SUVr by region. A. Brain regions with elevated PiB for those in the Early PiB group (n=1,088). For each specific brain region, the percentage of participants within the Early PiB group who had elevated PiB in respective regions by side is displayed. The brain regions are sorted high to low and shown as left (red square) and right (blue triangle) and also by voxel weighted median of the right and left hemisphere (black circle). B. Surface renderings of the percentage of participants with elevated PiB. Surface renderings of the percentage of participants with elevated PiB for each brain region is shown for each subgroup (very low, low, low-moderate, moderate, moderate-high, and high). Maps of both the left and the right hemispheres are shown for individual subgroups.

Estimation of a regional amyloid-beta progression by sub-grouping the participants using regional frequencies of amyloid-positivity revealed unique early patterns of amyloid burden in the brain (Figure 2B and supplementary figure 1 with detailed ROI data). The temporal, posterior cingulate, and occipital cortices and angular gyrus are seen to show early elevated PiB compared to other cortices in the ‘very low’ subgroup. Unique regional patterns appeared throughout the subgroups and eventually saturated all regions with elevated PiB-PET signal in the ‘high’ subgroup. Additionally, fusiform, inferior and the middle temporal region, middle temporal pole, posterior cingulate, angular gyrus, calcarine, and the inferior and the middle occipital lobe showed consistently elevated PiB-PET signal higher than the mean or regional percentage of other regions in the subgroups until all regions became saturated (Figure 2B and supplementary figure 1). Relationships between APOE genotype and early PiB SUVr were considered, however both APOE genotypes showed similar patterns visually (APOE ε4 carriers in red dot and non-carriers in blue dot in Supplementary Figure 1), implying little effect of the genotypes on the regional burden of amyloid-beta. The actual median regional SUVr values for each subgroup are also shown on surface renderings (Supplementary Figure 2).

### 3.2 Hierarchical clustering

To investigate heterogeneity of regional trends of early PiB SUVr deposition, cluster analysis was used. The hierarchical cluster analysis included the moderate, moderate-high, and high subgroups of our Early PiB group. Each cluster revealed distinct spatial patterns of Aβ deposition in the brain (Figure 3A and B): 1) frontal cluster (red circle) showed higher PiB-PET signal in the frontal lobe and lower in the occipital lobe, 2) occipitoparietal cluster (green triangles) showed higher PiB-PET signal in both the parietal and occipital lobes and lower in the frontal lobe, and 3) global cluster (blue square) showed generally lower PiB-PET signal and diffused patterns than the other two. Pair-wise statistical comparisons of the mean regional SUVr between clusters are shown in the Supplementary Figure 3 (Student’s two-sample t-test). The t-distributed stochastic neighbor embedding (TSNE) projection results also showed a distinct grouping between the clusters (Figure 3C). Particularly, the global PiB SUVr was not significantly different between frontal cluster vs. occipitoparietal cluster, however two clusters showed convincingly different PiB uptake level for frontal and occipitoparietal regions (lower panels in Figure 3C and Supplementary Figure 3).

**Figure 3.**
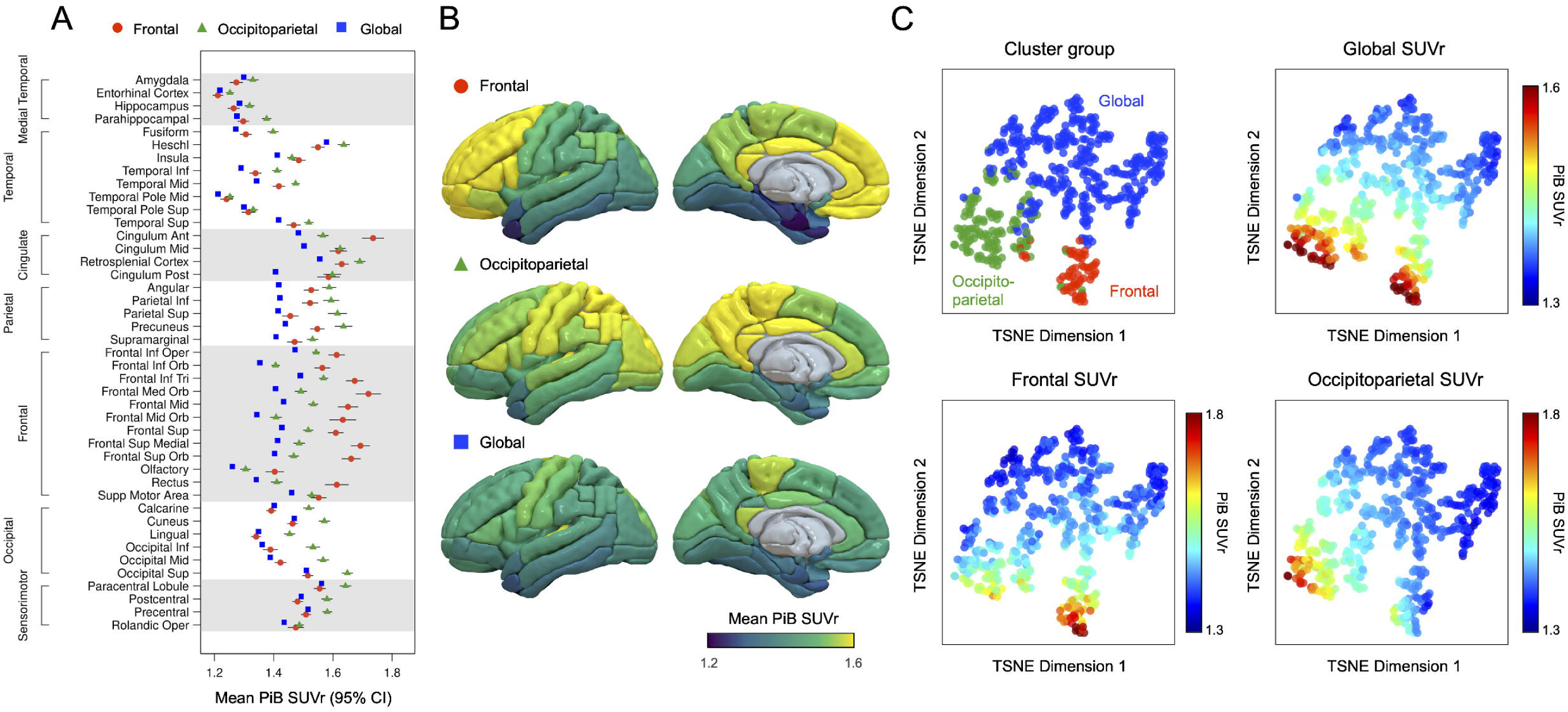
Hierarchical clustering analysis based on regional SUVr in the three highest subgroups (moderate, moderate-high, high) is shown. A. Regional mean PiB-PET SUVr is shown for each cluster (red circle: frontal cluster, green triangle: occipitoparietal cluster, and blue square: global cluster). Error bars indicate 95% confidence intervals. (B) 3D-rendering of mean SUVr map of each cluster. C. t-distributed stochastic neighbor embedding (TSNE) projection is illustrated with different colorcoding (i.e., cluster group, global SUVr, frontal SUVr, and occipitoparietal SUVr).

The clusters had unequal sizes, but were similar in diagnosis, age, and sex (table 2). APOE ε4 carriers were associated with the frontal and occipitoparietal cluster groups while non-carriers with the global cluster. In comparison of two types of cluster analysis, including K-means and hierarchical, both methods provided similar results. (see Supplementary figure 4). The hierarchical clustering (K=3) was performed using mean PiB SUVr over brain regions within each subgroup. Starting from low-moderate and moderate subgroup, a similar pattern of group separations (i.e., frontal, occipitoparietal and global) showing differences in parietal, frontal lobe and occipital lobe is observed (Supplementary figure 5).

### 3.4 Longitudinal changes of PiB-PET signals

To investigate the difference of degree of amyloid progression between clusters, annual PiB SUVr changes of participants with serial data in each cluster subgroups (n=33, 64, and 186 for the frontal cluster, occipitoparietal cluster and global cluster, respectively) were analyzed (Figure 4). The frontal group showed the highest amyloid-beta accumulation rates vs. other groups across the cortices followed by the occipitoparietal group.

**Figure 4.**
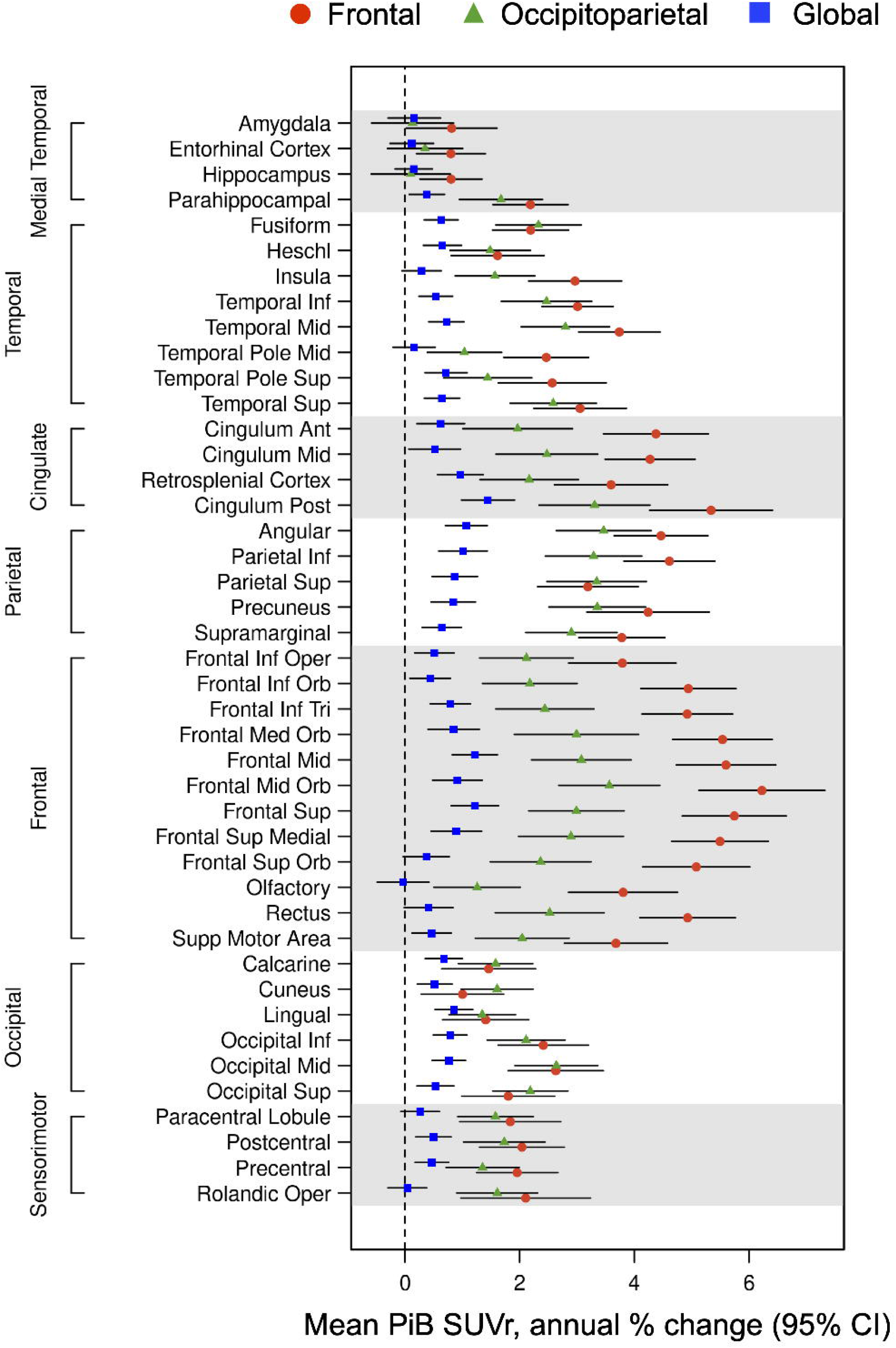
Annual PiB-PET SUVr change. Annual PiB-PET SUVr change was evaluated for individuals within the clusters who had serial data (n = 283). Error bars indicate 95% confidence intervals.

Comparing the frontal and the occipitoparietal groups, the frontal cluster showed a significantly higher accumulation rate in the frontal lobe and cingulate cortex (p<0.05, Student’s two-sample t-test; Supplementary Figure 6). The Occipital group also showed a higher progression compared to the global group (p<0.05, Student’s two-sample t-test; Supplementary Figure 6). The changes of cognitive test score (MMSE) and the clinical diagnosis were also considered; however, no significant difference was found among the cluster types.

## 4. DISCUSSION

This study revealed regional patterns of initial Aβ deposition within the neocortex. The use of region-specific cutoffs as determined in the young CN group allowed us to survey distinct areas that showed early Aβ distributions that may otherwise go unseen using traditional, global meta-ROI analysis. We showed that the earliest observed elevated PiB-PET signals were in the temporal, cingulate, and occipital regions. The percentage of those in each subgroup with elevated Aβ in these specific regions also increased sequentially with increasing global SUVr even when it was below typical global cut off thresholds. Other regions were identified that also showed a sequential elevated PiB-PET in relatively consistent patterns. We found that early regional Aβ patterns can be seen in both APOE carriers and non-carriers.

The initial areas of Aβ deposition seen included the temporal, cingulate, and occipital lobes. Namely, fusiform, inferior temporal lobe, middle temporal region, middle temporal pole, superior temporal lobe, posterior cingulum, angular gyrus, calcarine, cuneus, lingual, inferior occipital lobe, middle occipital lobe, and superior occipital lobe. Of these, the fusiform, angular gyrus inferior temporal, and the middle temporal region showed the greatest percentage of participants with elevated PiB levels in early patterns of deposition. Studies analyzing early deposition patterns of Aβ have found differing results, leading to large discrepancies of exactly where initial Aβ is accumulating. These discrepancies include initial aggregation sites found across the frontal lobe,[15-17] parietal,[16, 17] and temporal areas[14]; although others claim temporal areas are the latter points of aggregation.[15] These data suggest that in the earliest subgroups of Aβ accumulation, initial rise is seen in the temporal lobe, posterior cingulate region, and the occipital lobe. Additionally, we also showed that distinct early deposition patterns are apparent in different subgroups.

These findings are supported by theories of the functional connectivity and activity within the brain.[16] Both high neuronal connectivity and activity have been linked to the release and deposition of Aβ.[29, 30] The high neuronal connectivity of the posterior cingulate[31] as well as the occipital lobe[32, 33] appears to make these regions more vulnerable to Aβ deposition, as seen in our results and others.[30, 34] Our results showing early Aβ load in the middle prefrontal cortex, posterior cingulate, precuneus, and angular gyrus supports the idea that the default mode network (DMN) may relate with Aβ deposition.[16] The DMN includes brain regions with high connectivity, particularly in a spontaneous resting state[35] and has been shown to be vulnerable to Aβ deposition.[32, 36]

Late Aβ deposition in the sensorimotor cortex was also observed. Aβ load in this region has shown conflicting results in the past, with some claiming there is deposition in the sensorimotor cortex.[37] Our results found this region to have slower deposition rates across subgroups when compared to other regions (Figure 3), but steadily increasing SUVr values across the subgroups (Supplementary Figure 2). A possible explanation could be that the sensorimotor cortex is hyperexcitable,[29] giving higher susceptibility to Aβ deposition late in the disease, but possibly not at early stages.[34, 38] However, there is an lack of explanation as to why this area has the lowest Aβ deposition.[37]

We defined several subgroups with distinct patterns of early regional PiB-PET signal using clustering analysis. These included three distinct patterns of Aβ load in the brain: high in the frontal lobe and low in the parietal and occipital lobes (frontal cluster), high in the parietal and occipital lobes and lower in the frontal lobe (occipitoparietal cluster), and low in the temporal, parietal, frontal, and occipital lobes (global cluster). This observation aligns with a recent study that reported three sub-types of spatial-temporal amyloid accumulation (i.e., frontal, parietal and occipital).[39] The cingulate and sensorimotor cortices had similar levels of deposition between clusters. The parahippocampal gyrus, fusiform, inferior and the middle temporal region, and sensorimotor cortices showed higher Aβ load in the occipitoparietal cluster and the anterior cingulate cortex had higher Aβ deposition in the frontal cluster. Interestingly, the global cluster group showed similar regional frequencies of amyloid-positivity to other participants included in the analysis, but the global SUVr was significantly lower compared to other clusters (Figure 3C and Table 2). There is limited information about the heterogeneities in initial Aβ regional deposition; but it has been seen that regional prevalence of cerebral amyloid deposition differs across individuals- even for those already presenting cognitive impairment.[40] The clinical implications of these heterogeneities are not understood; however, their appearance in our results suggests early development of different subgroup-related phenotypes and future analysis and correlation with tau deposition patterns and clinical outcome is needed. Future work will involve further refining cluster groups when more participants can be evaluated.

APOE ε4 carriers made up 30.3%, 47.4%, and 20.7% of participants in the frontal, occipitoparietal and global cluster respectively. In the frontal and occipitoparietal clusters, where there was a higher percentage of participants who were APOE ε4 carriers, the parietal and frontal lobes had relatively higher PiB SUVr. Others have shown that APOE ε4 carriers have heightened levels of Aβ deposition in the frontal parietal regions, validating these patterns.[18] There were fewer APOE ε4 carriers in the global cluster, where deposition was low across multiple areas of the brain again suggesting that APOE carriers may have specific patterns of Aβ deposition within the brain that differ from non-carriers.

In the longitudinal analysis, the brain regions that showed a higher relative longitudinal Aβ progression includes the frontal, cingulate, temporal, parietal, and occipital lobes, consistent with the past studies.[11, 15, 17] The comparison between the subgroups showed that the frontal cluster had higher Aβ longitudinal deposition than others. The occipitoparietal group also showed higher rates of accumulation than the global cluster however, a lower annual percent change was seen in the frontal and cingulate cortices than the frontal cluster. The result aligns with the fact that being an APOE ε4 carrier heightens the risk of Aβ deposition[41] and causes its deposition earlier in life given the high proportion of APOE ε4 carriers in the frontal and occipitoparietal clusters.

Possible limitations of this study include that confirmation of these early PET findings is difficult. No cognitive abnormalities are generally present. Some studies suggest that cerebrospinal fluid (CSF) can detect abnormal Aβ before PET but autopsy confirmation is needed.[42] Lowered β-amyloid42 in CSF is strongly correlated with the presentation of early amyloid load in preclinical AD stages[42] and correlated to APOE carriers.[43] A possible comparison of early PET findings and CSF could be helpful. Additionally, our study does not have many AD dementia participants (n=0.6%). Therefore, we cannot confirm with these data that the patterns we observed are associated with eventual AD, even though this is a possible outcome for most. This is an area of current investigations. Despite this limitation, it is important to study Aβ deposition early, within CU individuals, given that Aβ deposition may begin before dementia occurs by ∼20 years.[44]

Our findings demonstrate that initial Aβ deposition occurs in specific brain regions and that some subgroups have distinct patterns of deposition that may represent different clinical phenotypes. In these distinct subgroups, amyloid deposition patterns are linked to APOE status. Although past studies have inconsistencies in describing early aggregation areas as described above, this may only be a demonstration of the presence of different subgroups in each study. We suggest that when larger cohorts are considered, the earliest patterns of Aβ are seen as a heterogeneous mix of pattern subtypes that represent different paths of Aβ deposition that may eventually predispose to distinct AD phenotypes. Identifying these regions of early aggregation and examining their properties in a population study may best elucidate how Aβ aggregation starts in sporadic AD. This knowledge is crucial in advancing both diagnostic techniques, understanding the development of AD phenotypes, and developing disease-modifying drugs.

### Sources of Funding

National Institutes of Health grant R01 AG073282 (V.L.), National Institutes of Health grant P30 AG62677-2 (D.J.), National Institutes of Health grant R01 AG011378 (C.J.), National Institutes of Health grant R01 AG041851 (C.J.), National Institutes of Health grant P50 AG016574 (R.P.), National Institutes of Health grant U01 AG06786 (R.P.), Robert Wood Johnson Foundation, The Elsie and Marvin Dekelboum Family Foundation, The Edson Family Foundation, The Liston Family Foundation The Robert H. and Clarice Smith and Abigail van Buren Alzheimer’s Disease Research Program, The GHR Foundation, Foundation Dr. Corinne Schuler (Geneva, Switzerland), Race Against Dementia, and the Mayo Foundation.

### Disclosures

Authors declare that they have no competing interests.

## Figure legends

**Table 1.** Subgroup demographics consisting of the Early PiB subgroups and the younger cognitively unimpaired group. The ANOVA and Pearson’s Chi-squared test indicates differences in age, education, diagnosis, and the global PiB SUVr value between the subgroups.

**Table 2.** Demographics of the cluster populations from Figure 3. The ANOVA and Pearson’s Chi-squared test indicates differences in APOE and global PiB SUVr value between the clusters.

**Supplementary Figure 1.**
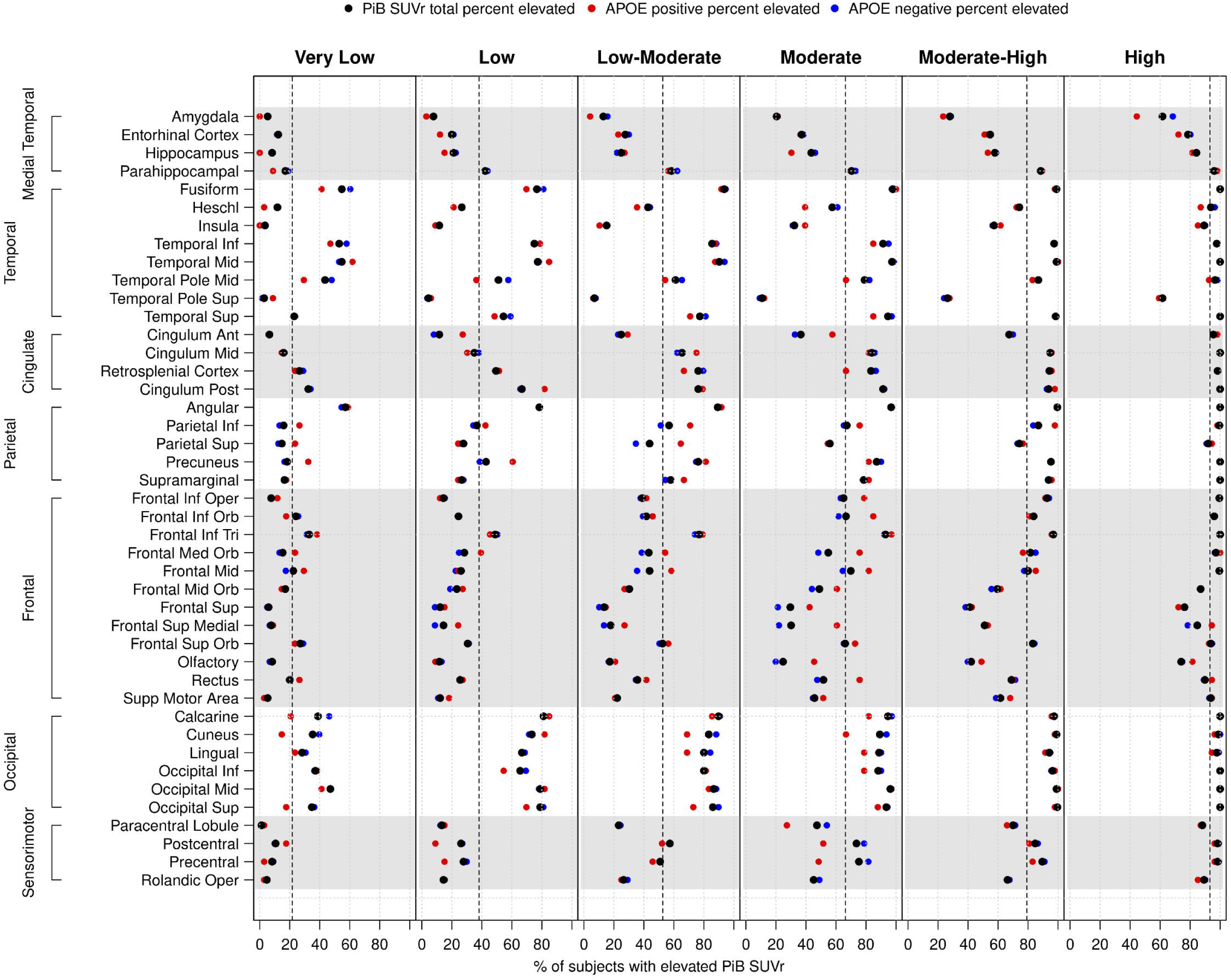
The percentage of participants in each subgroup with elevated PiB signal by brain region. The percentage of participants with elevated PiB for each brain region is shown (black dot) for each subgroup (very low, low, low-moderate, moderate, moderate-high, and high). Brain regions are grouped by lobe as indicated on the y-axis. The mean percentage of the number of regions with elevated global PiB for each subgroup is represented by a black dashed line and shows an increasing trend across subgroups as 20.26%, 37.66%, 53.11%, 66.31%, 80.06%, 93.41% from ‘very low’ to ‘high’. The red dot illustrates APOE carriers and the blue dot for APOE non-carriers.

**Supplementary Figure 2.**
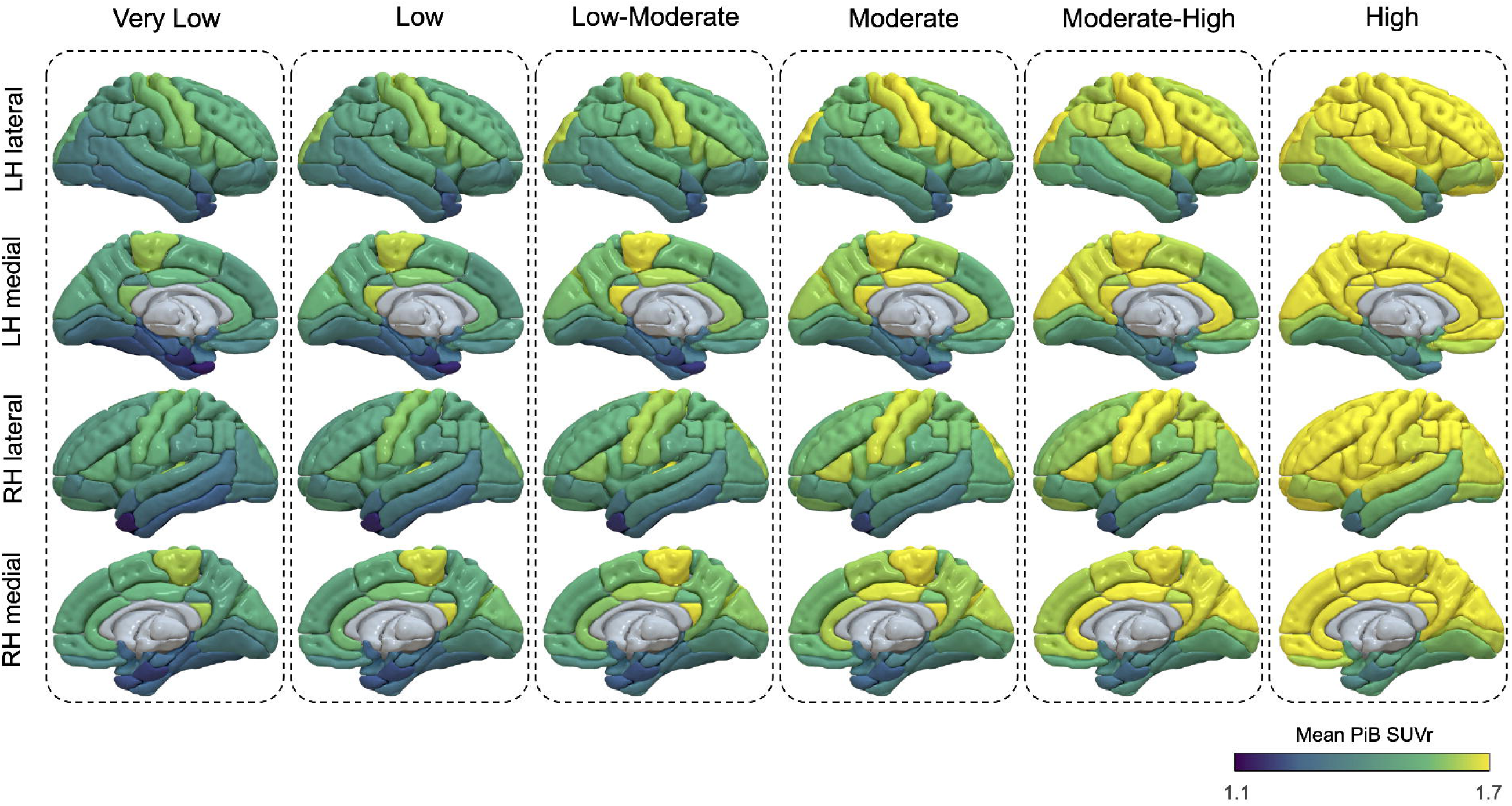
SUVr map of PiB displayed by brain regions in each subgroup.

**Supplementary Figure 3.**
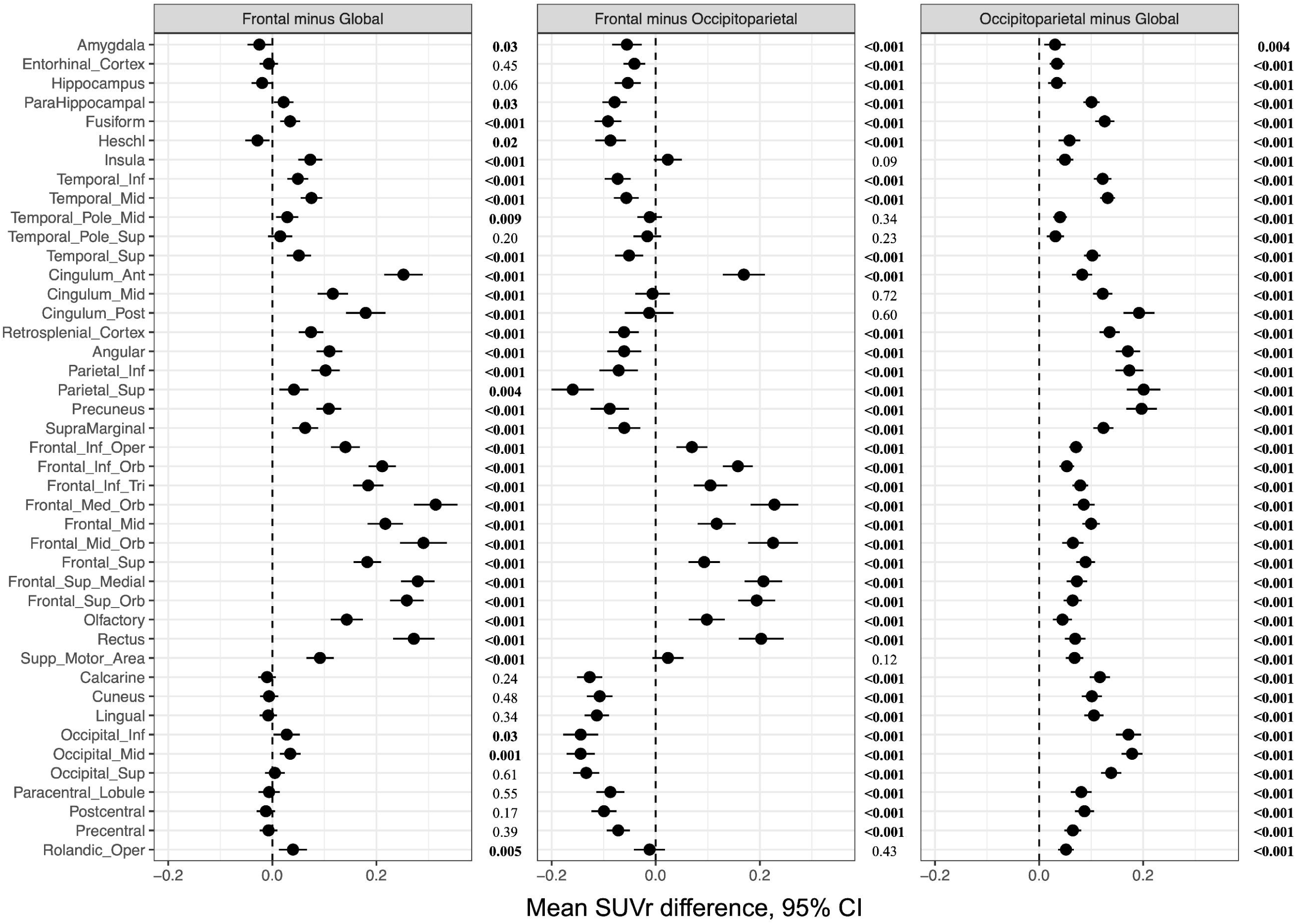
Pair-wise comparison of regional SUVr between clusters. The pair-wise comparisons of mean SUVr (i.e., frontal minus global, frontal minus occipitoparietal, and occipitoparietal minus global) were performed using a Student’s two-sample t-test. Error bars indicate 95% confidence intervals.

**Supplementary Figure 4.**
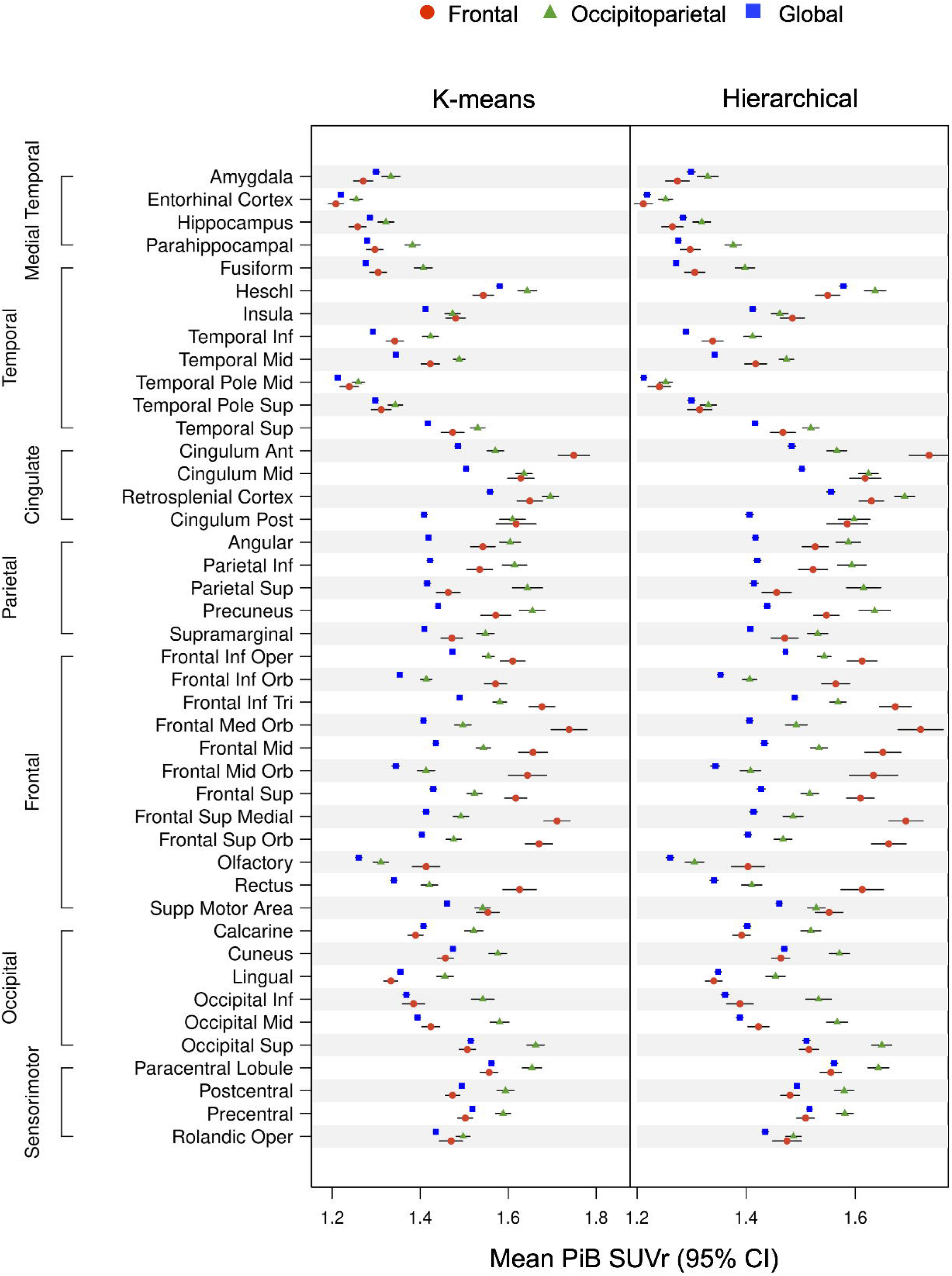
The comparison of two different clustering methods. K-mean clustering and hierarchical clustering, in the three highest subgroups of the Early PiB group (moderate, moderate-high, high) were compared. The number of clusters was restricted as 3 (K=3) for both K-mean (cluster 1; n=49, cluster 2; n=65, cluster 3; n=369) and hierarchical (cluster 1; n=30, cluster 2; n=36, cluster 3; n=417). Both algorithms showed similar results. Error bars indicate 95% confidence intervals.

**Supplementary Figure 5.**
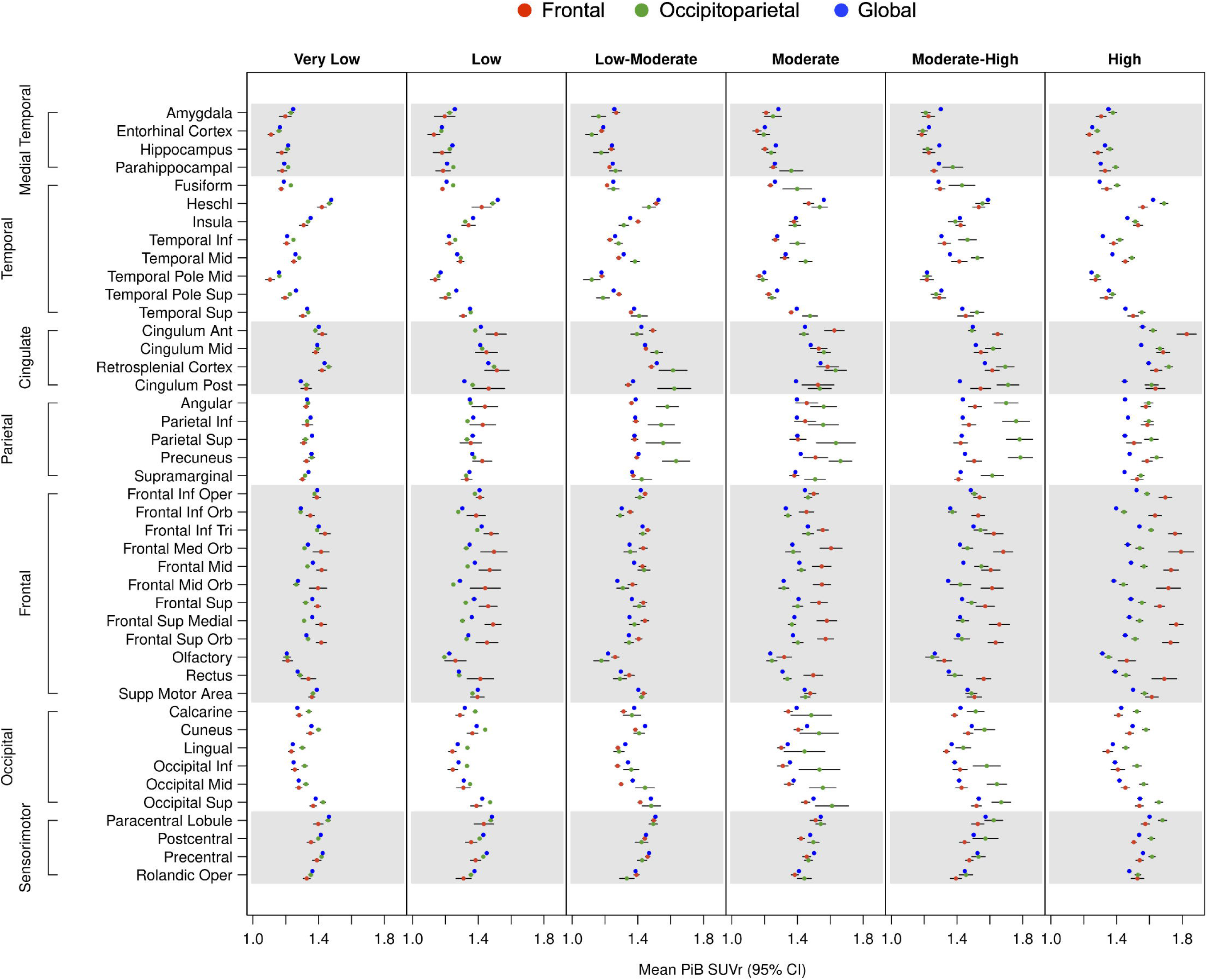
Regional PiB deposition in each subgroup by hierarchical cluster. Each column shows the clusters obtained with hierarchical clustering (K=3) using each subgroup. Clusters were analyzed by mean PiB SUVr over brain regions. Starting from low-moderate and moderate subgroup, a similar pattern showing differences in cingulate, frontal lobe and occipital lobe is observed.

**Supplementary Figure 6.**
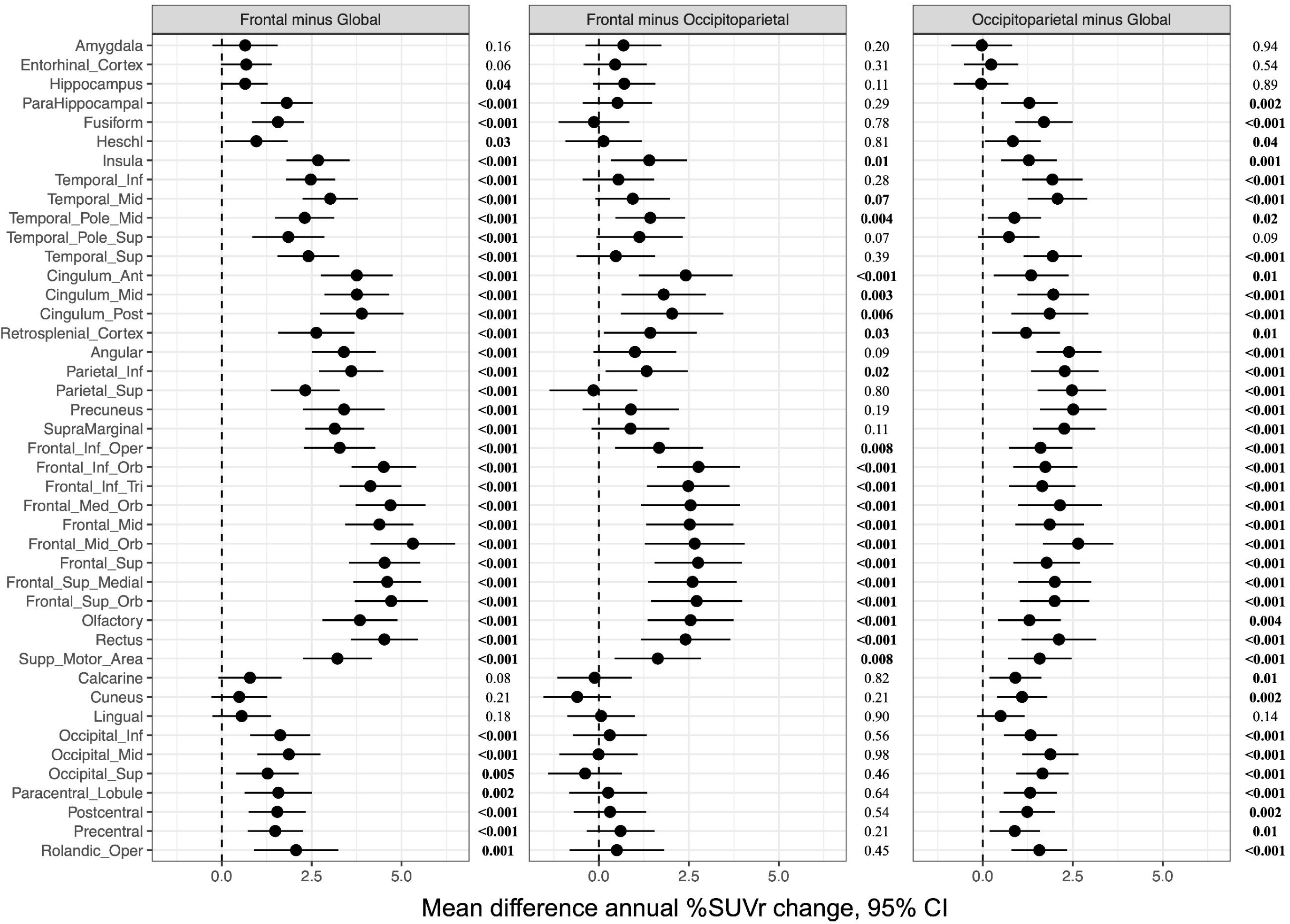
Pair-wise comparison of annual % SUVr change between clusters. The pair-wise comparisons of annual % SUVr change (i.e., frontal minus global, frontal minus occipitoparietal, and occipitoparietal minus global) were performed using a Student’s two-sample t-test.

**Supplementary Table 1.** Demographics for overall MCSA 50+ population, populations to compute selection criteria tertiles (MCSA 50+ CU, MCSA 50+ A+), and overall Early PiB population.

**Supplementary Table 2.** ROI specific SUVr cut points derived from younger cognitively unimpaired individuals in the MCSA (30-49 years, n=164). Each regional cut point value is from the 95th percentile per ROI of the younger cognitively unimpaired individuals. The cut points for the left hemisphere, right hemisphere, and bilateral brain were separately calculated for each brain region.

## Data Availability

Data may be available from the authors upon reasonable request and with permission.

## REFERENCES

[1] Crews L, Masliah E. Molecular mechanisms of neurodegeneration in Alzheimer’s disease. Human molecular genetics. 2010;19:R12–R20.

[2] Terry RD, Masliah E, Salmon DP, Butters N, DeTeresa R, Hill R, et al. Physical basis of cognitive alterations in Alzheimer’s disease: synapse loss is the major correlate of cognitive impairment. Annals of Neurology: Official Journal of the American Neurological Association and the Child Neurology Society. 1991;30:572–80.

[3] Klunk WE, Engler H, Nordberg A, Wang Y, Blomqvist G, Holt DP, et al. Imaging brain amyloid in Alzheimer’s disease with Pittsburgh CompoundLJB. Annals of Neurology: Official Journal of the American Neurological Association and the Child Neurology Society. 2004;55:306–19.

[4] Zhang S, Han D, Tan X, Feng J, Guo Y, Ding Y. Diagnostic accuracy of 18FLJFDG and 11CLJPIBLJPET for prediction of shortLJterm conversion to Alzheimer’s disease in subjects with mild cognitive impairment. International journal of clinical practice. 2012;66:185–98.

[5] Cohen AD, Landau SM, Snitz BE, Klunk WE, Blennow K, Zetterberg H. Fluid and PET biomarkers for amyloid pathology in Alzheimer’s disease. Molecular and cellular neuroscience. 2019;97:3–17.

[6] Lowe VJ, Lundt E, Knopman D, Senjem ML, Gunter JL, Schwarz CG, et al. Comparison of [18F] Flutemetamol and [11C] Pittsburgh Compound-B in cognitively normal young, cognitively normal elderly, and Alzheimer’s disease dementia individuals. NeuroImage: Clinical. 2017;16:295–302.

[7] Wolk DA, Grachev ID, Buckley C, Kazi H, Grady MS, Trojanowski JQ, et al. Association between in vivo fluorine 18–labeled flutemetamol amyloid positron emission tomography imaging and in vivo cerebral cortical histopathology. Archives of neurology. 2011;68:1398–403.

[8] Byun MS, Yoon Y, Kim G, Yi D, Shin SA, Kim YK, et al. O2LJ03LJ06: HETEROGENEITY OF AMYLOID DEPOSITION PATTERN AMONG AMYLOIDLJPOSITIVE COGNITIVELY IMPAIRED INDIVIDUALS. Alzheimer’s & Dementia. 2019;15:P542-P.

[9] Engler H, Forsberg A, Almkvist O, Blomquist G, Larsson E, Savitcheva I, et al. Two-year follow-up of amyloid deposition in patients with Alzheimer’s disease. Brain. 2006;129:2856–66.

[10] Grimmer T, Tholen S, Yousefi BH, Alexopoulos P, Förschler A, Förstl H, et al. Progression of cerebral amyloid load is associated with the apolipoprotein E ε4 genotype in Alzheimer’s disease. Biological psychiatry. 2010;68:879–84.

[11] Villemagne VL, Pike KE, Chételat G, Ellis KA, Mulligan RS, Bourgeat P, et al. Longitudinal assessment of Aβ and cognition in aging and Alzheimer disease. Annals of neurology. 2011;69:181–92.

[12] Thal DR, Rüb U, Orantes M, Braak H. Phases of Aβ-deposition in the human brain and its relevance for the development of AD. Neurology. 2002;58:1791–800.

[13] Bharadwaj PR, Dubey AK, Masters CL, Martins RN, Macreadie IG. Aβ aggregation and possible implications in Alzheimer’s disease pathogenesis. Journal of cellular and molecular medicine. 2009;13:412–21.

[14] Cho H, Choi JY, Hwang MS, Kim YJ, Lee HM, Lee HS, et al. In vivo cortical spreading pattern of tau and amyloid in the Alzheimer disease spectrum. Annals of neurology. 2016;80:247–58.

[15] Grothe MJ, Barthel H, Sepulcre J, Dyrba M, Sabri O, Teipel SJ, et al. In vivo staging of regional amyloid deposition. Neurology. 2017;89:2031–8.

[16] Palmqvist S, Schöll M, Strandberg O, Mattsson N, Stomrud E, Zetterberg H, et al. Earliest accumulation of β-amyloid occurs within the default-mode network and concurrently affects brain connectivity. Nature communications. 2017;8:1214.

[17] Mattsson N, Palmqvist S, Stomrud E, Vogel J, Hansson O. Staging β-amyloid pathology with amyloid positron emission tomography. JAMA neurology. 2019;76:1319–29.

[18] Pletnikova O, Kageyama Y, Rudow G, LaClair KD, Albert M, Crain BJ, et al. The spectrum of preclinical Alzheimer’s disease pathology and its modulation by ApoE genotype. Neurobiology of aging. 2018;71:72–80.

[19] Jack Jr CR, Wiste HJ, Weigand SD, Therneau TM, Lowe VJ, Knopman DS, et al. Defining imaging biomarker cut points for brain aging and Alzheimer’s disease. Alzheimer’s & Dementia. 2017;13:205–16.

[20] Lowe VJ, Bruinsma TJ, Min H-K, Lundt ES, Fang P, Senjem ML, et al. Elevated medial temporal lobe and pervasive brain tau-PET signal in normal participants. Alzheimer’s & Dementia: Diagnosis, Assessment & Disease Monitoring. 2018;10:210–6.

[21] Roberts RO, Geda YE, Knopman DS, Cha RH, Pankratz VS, Boeve BF, et al. The Mayo Clinic Study of Aging: design and sampling, participation, baseline measures and sample characteristics. Neuroepidemiology. 2008;30:58–69.

[22] Petersen RC. Mild cognitive impairment as a diagnostic entity. Journal of internal medicine. 2004;256:183–94.

[23] Lowe VJ, Lundt ES, Senjem ML, Schwarz CG, Min H-K, Przybelski SA, et al. White matter reference region in PET studies of 11C-Pittsburgh compound B uptake: effects of age and amyloid-β deposition. Journal of Nuclear Medicine. 2018;59:1583–9.

[24] Tzourio-Mazoyer N, Landeau B, Papathanassiou D, Crivello F, Etard O, Delcroix N, et al. Automated anatomical labeling of activations in SPM using a macroscopic anatomical parcellation of the MNI MRI single-subject brain. Neuroimage. 2002;15:273–89.

[25] Lowe VJ, Wiste HJ, Senjem ML, Weigand SD, Therneau TM, Boeve BF, et al. Widespread brain tau and its association with ageing, Braak stage and Alzheimer’s dementia. Brain. 2018;141:271–87.

[26] Meltzer CC, Leal JP, Mayberg HS, Wagner Jr HN, Frost JJ. Correction of PET data for partial volume effects in human cerebral cortex by MR imaging. Journal of computer assisted tomography. 1990;14:561–70.

[27] Everitt BS, Landau S, Leese M, Stahl D. An introduction to classification and clustering. Cluster analysis. 2011;5:1–13.

[28] Lloyd S. Least squares quantization in PCM. IEEE transactions on information theory. 1982;28:129–37.

[29] Ferreri F, Vecchio F, Vollero L, Guerra A, Petrichella S, Ponzo D, et al. Sensorimotor cortex excitability and connectivity in Alzheimer’s disease: A TMSLJEEG coLJregistration study. Human brain mapping. 2016;37:2083–96.

[30] Li X, Uemura K, Hashimoto T, Nasser-Ghodsi N, Arimon M, Lill CM, et al. Neuronal activity and secreted amyloid β lead to altered amyloid β precursor protein and presenilin 1 interactions. Neurobiology of disease. 2013;50:127–34.

[31] Buckner RL, Sepulcre J, Talukdar T, Krienen FM, Liu H, Hedden T, et al. Cortical hubs revealed by intrinsic functional connectivity: mapping, assessment of stability, and relation to Alzheimer’s disease. Journal of neuroscience. 2009;29:1860–73.

[32] Hafkemeijer A, van der Grond J, Rombouts SA. Imaging the default mode network in aging and dementia. Biochimica et Biophysica Acta (BBA)-Molecular Basis of Disease. 2012;1822:431–41.

[33] Zhang H-Y, Wang S-J, Liu B, Ma Z-L, Yang M, Zhang Z-J, et al. Resting brain connectivity: changes during the progress of Alzheimer disease. Radiology. 2010;256:598–606.

[34] Cirrito JR, Kang J-E, Lee J, Stewart FR, Verges DK, Silverio LM, et al. Endocytosis is required for synaptic activity-dependent release of amyloid-β in vivo. Neuron. 2008;58:42–51.

[35] Mohan A, Roberto AJ, Mohan A, Lorenzo A, Jones K, Carney MJ, et al. Focus: the aging brain: the significance of the default mode network (DMN) in neurological and neuropsychiatric disorders: a review. The Yale journal of biology and medicine. 2016;89:49.

[36] Bero AW, Yan P, Roh JH, Cirrito JR, Stewart FR, Raichle ME, et al. Neuronal activity regulates the regional vulnerability to amyloid-β deposition. Nature neuroscience. 2011;14:750–6.

[37] Fantoni E, Collij L, Alves IL, Buckley C, Farrar G. The spatial-temporal ordering of amyloid pathology and opportunities for PET imaging. Journal of Nuclear Medicine. 2020;61:166–71.

[38] Bero AW, Bauer AQ, Stewart FR, White BR, Cirrito JR, Raichle ME, et al. Bidirectional relationship between functional connectivity and amyloid-β deposition in mouse brain. Journal of neuroscience. 2012;32:4334–40.

[39] Collij LE, Salvadó G, Wottschel V, Mastenbroek SE, Schoenmakers P, Heeman F, et al. Spatial-temporal patterns of β-amyloid accumulation: a subtype and stage inference model analysis. Neurology. 2022;98:e1692–e703.

[40] Byun MS, Kim SE, Park J, Yi D, Choe YM, Sohn BK, et al. Heterogeneity of regional brain atrophy patterns associated with distinct progression rates in Alzheimer’s disease. PLoS One. 2015;10:e0142756.

[41] Rodrigue KM, Kennedy KM, Park DC. Beta-amyloid deposition and the aging brain. Neuropsychology review. 2009;19:436–50.

[42] Palmqvist S, Mattsson N, Hansson O, Initiative AsDN. Cerebrospinal fluid analysis detects cerebral amyloid-β accumulation earlier than positron emission tomography. Brain. 2016;139:1226–36.

[43] Mattsson N, Insel PS, Donohue M, Landau S, Jagust WJ, Shaw LM, et al. Independent information from cerebrospinal fluid amyloid-β and florbetapir imaging in Alzheimer’s disease. Brain. 2015;138:772–83.

[44] Bateman RJ, Xiong C, Benzinger TL, Fagan AM, Goate A, Fox NC, et al. Clinical and biomarker changes in dominantly inherited Alzheimer’s disease. N Engl J Med. 2012;367:795–804.

