## Supplementary material for "Patterns of early neocortical amyloid beta accumulation: a positron emission tomography population-based study": Table

|  | Younger  cognitively  unimpaired (N=164) | Very Low (N=170) | Low (N=180) | Low-Moderate (N=185) | Moderate (N=186) | Moderate-High (N=190) | High (N=177) | p value |
| --- | --- | --- | --- | --- | --- | --- | --- | --- |
| **Age, years** |  |  |  |  |  |  |  | < 0.001^1^ |
| Mean (SD) | 41 (6) | 67 (10) | 71 (10) | 72 (9) | 74 (9) | 75 (9) | 75 (8) |  |
| Range | 31 - 50 | 51 - 92 | 50 - 94 | 52 - 93 | 52 - 91 | 54 - 93 | 53 - 93 |  |
| **Sex, N (%)** |  |  |  |  |  |  |  | 0.597^2^ |
| Female | 73 (44.5%) | 85 (50.0%) | 80 (44.4%) | 87 (47.0%) | 99 (53.2%) | 95 (50.0%) | 82 (46.3%) |  |
| Male | 91 (55.5%) | 85 (50.0%) | 100 (55.6%) | 98 (53.0%) | 87 (46.8%) | 95 (50.0%) | 95 (53.7%) |  |
| **Education, years** |  |  |  |  |  |  |  | < 0.001^1^ |
| N-Miss | 0 | 0 | 1 | 0 | 0 | 0 | 0 |  |
| Mean (SD) | 15.76 (2.24) | 15.25 (2.62) | 14.84 (2.73) | 14.55 (2.55) | 14.30 (2.56) | 14.33 (2.74) | 14.37 (2.68) |  |
| Range | 11.00 - 20.00 | 9.00 - 20.00 | 7.00 - 20.00 | 8.00 - 20.00 | 7.00 - 20.00 | 0.00 - 20.00 | 6.00 - 20.00 |  |
| **Diagnosis, N (%)** |  |  |  |  |  |  |  |  |
| N-Miss | 0 | 1 | 1 | 1 | 0 | 1 | 2 |  |
| CU | 164 (100.0%) | 159 (94.1%) | 162 (90.5%) | 168 (91.3%) | 165 (88.7%) | 165 (87.3%) | 150 (85.7%) |  |
| MCI | 0 (0.0%) | 10 (5.9%) | 17 (9.5%) | 15 (8.2%) | 20 (10.8%) | 22 (11.6%) | 23 (13.1%) |  |
| DEM | 0 (0.0%) | 0 (0.0%) | 0 (0.0%) | 1 (0.5%) | 1 (0.5%) | 2 (1.1%) | 2 (1.1%) |  |
| OTHER | 0 (0.0%) | 0 (0.0%) | 0 (0.0%) | 0 (0.0%) | 0 (0.0%) | 0 (0.0%) | 0 (0.0%) |  |
| **APOE ε4, N (%)** |  |  |  |  |  |  |  | 0.040^2^ |
| N-Miss | 19 | 15 | 10 | 10 | 12 | 10 | 12 |  |
| Non-carrier | 112 (77.2%) | 121 (78.1%) | 137 (80.6%) | 127 (72.6%) | 141 (81.0%) | 133 (73.9%) | 111 (67.3%) |  |
| Carrier | 33 (22.8%) | 34 (21.9%) | 33 (19.4%) | 48 (27.4%) | 33 (19.0%) | 47 (26.1%) | 54 (32.7%) |  |
| **GM PVC PiB SUVr** |  |  |  |  |  |  |  | < 0.001^1^ |
| Mean (SD) | 1.23 (0.05) | 1.31 (0.01) | 1.33 (0.02) | 1.35 (0.02) | 1.38 (0.04) | 1.42 (0.05) | 1.49 (0.07) |  |
| Range | 1.10 - 1.40 | 1.30 - 1.35 | 1.30 - 1.44 | 1.31 - 1.46 | 1.33 - 1.58 | 1.36 - 1.62 | 1.39 - 1.62 |  |

**Table 1.** Subgroup demographics consisting of the Early PiB subgroups and the younger cognitively unimpaired group. The ANOVA and Pearson’s Chi-squared test indicates differences in age, education, diagnosis, and the global PiB SUVr value between the subgroups.

**Table 2.** Demographics of the cluster populations from Figure 3. The ANOVA and Pearson’s Chi-squared test indicates differences in APOE and global PiB SUVr value between the clusters.

|  | Frontal (N=131) | Occipitoparietal (N=60) | Global (N=362) | p value |
| --- | --- | --- | --- | --- |
| **Age, years** |  |  |  | < 0.001^1^ |
| Mean (SD) | 78 (8) | 75 (7) | 73 (8) |  |
| Range | 53 - 93 | 61 - 90 | 52 - 93 |  |
| **Sex, N (%)** |  |  |  | 0.035^2^ |
| Female | 53 (40.5%) | 29 (48.3%) | 194 (53.6%) |  |
| Male | 78 (59.5%) | 31 (51.7%) | 168 (46.4%) |  |
| **Education, years** |  |  |  | 0.476^1^ |
| Mean (SD) | 14.58 (2.60) | 14.27 (2.98) | 14.25 (2.62) |  |
| Range | 7.00 - 20.00 | 6.00 - 20.00 | 0.00 - 20.00 |  |
| **Diagnosis, N (%)** |  |  |  | 0.761^2^ |
| N-Miss | 1 | 0 | 2 |  |
| 0=CU | 111 (85.4%) | 53 (88.3%) | 316 (87.8%) |  |
| 1=MCI | 17 (13.1%) | 6 (10.0%) | 42 (11.7%) |  |
| 3=Dementia | 2 (1.5%) | 1 (1.7%) | 2 (0.6%) |  |
| **APOE ε4, N (%)** |  |  |  | < 0.001^2^ |
| N-Miss | 12 | 3 | 19 |  |
| Non-carrier | 83 (69.7%) | 30 (52.6%) | 272 (79.3%) |  |
| Carrier | 36 (30.3%) | 27 (47.4%) | 71 (20.7%) |  |
| **GM PVC PiB SUVr** |  |  |  | < 0.001^1^ |
| Mean (SD) | 1.50 (0.06) | 1.52 (0.06) | 1.39 (0.03) |  |
| Range | 1.37 - 1.62 | 1.42 - 1.62 | 1.33 - 1.49 |  |

**Supplementary Table 1.** Demographics for overall MCSA 50+ population, populations to compute selection criteria tertiles (MCSA 50+ CU, MCSA 50+ A+), and overall Early PiB population.

|  | MCSA 50+ (N=2255) | MCSA 50+ CU (N=1947) | MCSA 50+ A+ (N=703) | Early PiB (N=1088) |
| --- | --- | --- | --- | --- |
| **Age, years** |  |  |  |  |
| Mean (SD) | 72 (10) | 70 (10) | 78 (8) | 72 (9) |
| Range | 50 - 95 | 50 - 95 | 53 - 95 | 50 - 94 |
| **Sex, N (%)** |  |  |  |  |
| Female | 1062 (47.1%) | 933 (47.9%) | 336 (47.8%) | 528 (48.5%) |
| Male | 1193 (52.9%) | 1014 (52.1%) | 367 (52.2%) | 560 (51.5%) |
| **Education, years** |  |  |  |  |
| N-Miss | 1 | 0 | 0 | 1 |
| Mean (SD) | 14.77 (2.66) | 14.95 (2.54) | 14.54 (2.72) | 14.60 (2.66) |
| Range | 0.00 - 20.00 | 6.00 - 20.00 | 6.00 - 20.00 | 0.00 - 20.00 |
| **Diagnosis, N (%)** |  |  |  |  |
| N-Miss | 7 | 0 | 2 | 6 |
| CU | 1947 (86.6%) | 1947 (100.0%) | 530 (75.6%) | 969 (89.6%) |
| MCI | 265 (11.8%) | 0 (0.0%) | 141 (20.1%) | 107 (9.9%) |
| DEM | 32 (1.4%) | 0 (0.0%) | 28 (4.0%) | 6 (0.6%) |
| OTHER | 4 (0.2%) | 0 (0.0%) | 2 (0.3%) | 0 (0.0%) |
| **APOE ε4, N (%)** |  |  |  |  |
| N-Miss | 134 | 118 | 40 | 69 |
| Non-carrier | 1505 (71.0%) | 1327 (72.6%) | 356 (53.7%) | 770 (75.6%) |
| Carrier | 616 (29.0%) | 502 (27.4%) | 307 (46.3%) | 249 (24.4%) |
| **GM PVC PiB SUVr** |  |  |  |  |
| Mean (SD) | 1.51 (0.42) | 1.46 (0.35) | 1.97 (0.49) | 1.38 (0.07) |
| Range | 1.08 - 3.85 | 1.08 - 3.36 | 1.42 - 3.85 | 1.30 - 1.62 |

**Supplementary Table 2.** ROI specific SUVr cut points derived from younger cognitively unimpaired individuals in the MCSA (30-49 years, n=164). Each regional cut point value is from the 95th percentile per ROI of the younger cognitively unimpaired individuals. The cut points for the left hemisphere, right hemisphere, and total brain were separately calculated for each brain region.

| Lobe | Region | PiB SUVr left | PiB SUVr right | PiB SUVr total |
| --- | --- | --- | --- | --- |
| Medial Temporal | Amygdala | 1.38 | 1.33 | 1.33 |
|  | Entorhinal Cortex | 1.23 | 1.21 | 1.21 |
|  | Hippocampus | 1.28 | 1.28 | 1.27 |
|  | Parahippocampal | 1.25 | 1.24 | 1.23 |
| Temporal | Fusiform | 1.18 | 1.21 | 1.19 |
|  | Heschl | 1.52 | 1.56 | 1.53 |
|  | Insula | 1.41 | 1.43 | 1.41 |
|  | Temporal Inf | 1.19 | 1.24 | 1.22 |
|  | Temporal Mid | 1.24 | 1.28 | 1.26 |
|  | Temporal Pole Mid | 1.15 | 1.21 | 1.16 |
|  | Temporal Pole Sup | 1.32 | 1.34 | 1.33 |
|  | Temporal Sup | 1.33 | 1.37 | 1.35 |
| Cingulate | Cingulum Ant | 1.48 | 1.46 | 1.47 |
|  | Cingulum Mid | 1.42 | 1.44 | 1.43 |
|  | Retrosplenial Cortex | 1.44 | 1.52 | 1.47 |
|  | Cingulum Post | 1.31 | 1.33 | 1.32 |
| Parietal | Angular | 1.34 | 1.31 | 1.32 |
|  | Parietal Inf | 1.40 | 1.36 | 1.37 |
|  | Parietal Sup | 1.40 | 1.38 | 1.39 |
|  | Precuneus | 1.37 | 1.37 | 1.38 |
|  | Supramarginal | 1.35 | 1.38 | 1.36 |
| Frontal | Frontal Inf Oper | 1.40 | 1.46 | 1.43 |
|  | Frontal Inf Orb | 1.33 | 1.32 | 1.32 |
|  | Frontal Inf Tri | 1.42 | 1.45 | 1.41 |
|  | Frontal Med Orb | 1.35 | 1.41 | 1.37 |
|  | Frontal Mid | 1.37 | 1.42 | 1.39 |
|  | Frontal Mid Orb | 1.32 | 1.33 | 1.33 |
|  | Frontal Sup | 1.43 | 1.47 | 1.45 |
|  | Frontal Sup Medial | 1.42 | 1.45 | 1.42 |
|  | Frontal Sup Orb | 1.41 | 1.35 | 1.36 |
|  | Olfactory | 1.29 | 1.31 | 1.28 |
|  | Rectus | 1.33 | 1.32 | 1.33 |
|  | Supp Motor Area | 1.44 | 1.45 | 1.45 |
| Occipital | Calcarine | 1.30 | 1.29 | 1.29 |
|  | Cuneus | 1.38 | 1.38 | 1.38 |
|  | Lingual | 1.27 | 1.27 | 1.27 |
|  | Occipital Inf | 1.27 | 1.27 | 1.27 |
|  | Occipital Mid | 1.31 | 1.27 | 1.29 |
|  | Occipital Sup | 1.41 | 1.40 | 1.40 |
| Sensorimotor | Paracentral Lobule | 1.54 | 1.54 | 1.54 |
|  | Postcentral | 1.43 | 1.47 | 1.45 |
|  | Precentral | 1.46 | 1.48 | 1.46 |
|  | Rolandic Oper | 1.41 | 1.44 | 1.42 |
